# Regulation of Medical Devices in Zimbabwe: A qualitative study with key stakeholders

**DOI:** 10.1101/2023.06.07.23291092

**Authors:** Charles Chiku, Talkmore Maruta, Fredrick Mbiba, Justen Manasa

## Abstract

**Introduction:** Medical devices play a crucial role in promoting public health. However, there is a disparity in the availability and readiness of medical device regulations in Resource-Limited Settings (RLS) compared to the global average. We conducted a study to explore the medical device regulation landscape and stakeholder perceptions in Zimbabwe.

**Methodology:** Between June and November 2022, we administered questionnaires to representatives of the Medicines Control Authority of Zimbabwe (MCAZ), the Medical Laboratory and Clinical Scientists Council of Zimbabwe (MLCScCZ), and the National Microbiology Reference Laboratory (NMRL). We also conducted semi-structured interviews with 12 national-level critical stakeholders from these institutions to understand the current status of medical device regulations and the relationships between them. Additionally, we interviewed regulators from the South African Health Products Regulatory Authority, Tanzania, and World Health Organisation to learn about best practices in transitioning to medical device regulations. We used a thematic approach and an inductively developed common coding framework to analyse emerging themes.

**Results:** Our findings indicate that the current legal framework needs to be revised to regulate medical devices effectively, as it does not specify the institution(s) responsible for regulating them. While the MCAZ regulates condoms and gloves, the MLCScCZ coordinates with the NMRL to register In Vitro Diagnostic Medical Devices for priority diseases. Other medical devices are not regulated. Furthermore, conformity assessments for product registration are not proportional to the risk classification of medical devices, and post-market surveillance activities are ineffective. Stakeholders recognise the need to collaborate and improve the regulation of medical devices.

**Conclusion:** Zimbabwe must improve its regulatory framework for medical devices to ensure that safe medical devices of acceptable quality and performance are accessible. A solid legal foundation is necessary for harmonisation and reliance practices to reduce the regulatory burden on economic operators and ease the workload on regulators.

## Introduction

The medical device regulations are a set of interlinked legislation and regulations that govern the clinical investigation, production and distribution of medical devices, considering the medical device’s lifecycle based on its design, code of action and risk classification (1,2). A medical device is *"any instrument, apparatus, machine, appliance, implant, reagent for in vitro use, software, material, or related article used for a specific medical purpose"* (3). An in vitro device (IVD) is a medical device used to examine specimens collected from humans for diagnostic information, monitoring, or compatibility purposes (4). The general principle of medical devices regulation is to establish and implement controls intended to safeguard the health and safety of patients, users and other persons by ensuring that manufacturers of medical devices follow specified essential principles of safety and performance during design, manufacture and marketing. Additionally, the risk controls must be proportional to the risk classification of the medical device.

On the other hand, the regulatory controls must not impose an unnecessary regulatory burden on the economic operators (manufacturers, importers, authorised representatives, distributors and wholesalers) (5,6). Conformity assessment is based on the medical devices meeting all the applicable Essential Principles (EP), which will be demonstrated and assessed according to procedures designated by the Regulatory Authority and described in other Global Harmonisation Task Force (GHTF) and International Medical Devices Regulators Forum (IMDRF) guidances (7). The GHTF, which later changed to IMDRF, encourages and supports the global convergence of regulatory systems.

There are disparities in the availability of medical device regulatory services. According to the World Health Organisation (WHO), about 40% of countries in the WHO Africa (Afro) region have no regulations for medical devices, 32% have some regulations, and 28% have no data. In contrast, at the global level, 58% of all WHO member states indicated they had medical device regulations (8). This gap in medical device regulation between the Afro region and the global average may translate to lower-quality medical devices and hinder access to quality-assured medical devices.

Zimbabwe is a low-income country in Southern Africa that may be affected by the disparity of medical device regulation. In a study conducted by Hubner et al., it was concluded that Zimbabwe had a legal framework for regulating medical devices, conformity assessment (evaluation conducted to approve medical devices to be granted market access), import and export and post-market surveillance for condoms and gloves only (9). A qualitative study to assess the regulation of HIV-Self Testing IVDs in Malawi, Zambia and Zimbabwe was conducted by Dacombe et al. It was determined that the mandate to regulate HIV-Self Testing IVDs overlapped between the Medical Laboratory and Clinical Scientists Council of Zimbabwe and the Medicines Control Authority of Zimbabwe (MCAZ). Stakeholders indicated they needed a better understanding of the process and requirements for HIVST regulation and more clarity and coordination between organisational roles (10).

Additionally, the non-existing or weak regulatory system may have economic costs on the country due to Substandard and Falsified (SF) medical devices. Underdeveloped regulatory systems are associated with the regulatory burden for manufacturers to comply with unclear or different regulations when seeking premarket approvals in other countries. Therefore, some manufacturers may be reluctant to introduce their medical devices in countries with underdeveloped regulatory systems, which makes it difficult for the population to access much-needed medical devices. On the other hand, the respective National Regulatory Authorities (NRAs) must balance ensuring that the NRAs protect the public’s health and users through approved, safe, effective, acceptable quality and performance medical devices while ensuring the efficient introduction of needed IVDs (6). There needs to be more data regarding medical device regulation specific to Zimbabwe. Therefore, we conducted a qualitative study to determine the current regulatory status of medical devices, including IVDs, to document the perceptions and suggestions of key stakeholders to propose a roadmap to strengthen medical device regulation.

## Methods

Between June and November 2022, we conducted in-depth interviews to understand stakeholders’ relationships and perceptions of current and future medical device regulation (11). Specifically, we sought to document stakeholders’ current understanding and knowledge of medical device regulations and to explore sensitive areas around how the development of regulation for medical devices can be influenced by context and individual stakeholders. Data were collected using two approaches. First, structured questionnaires containing closed and open-ended questions were administered to key informants purposively selected due to their critical skills and knowledge of medical device regulations. With this, we aimed to get an overview of the premarket and post-market controls in place to ensure that only medical devices that were safe and effective were granted market authorisation and monitored after approval. Questions were adopted from the study conducted by Rugera et al. that sought to determine the regulation of medical devices in the East African Community partner states. Key areas addressed were (i) the existence and role of regulatory authorities; (ii) policy and legal framework for regulation; (iii) premarket control; (iv) marketing controls; (v) post-marketing controls(12). The tool was composed of both open and closed-ended questions relating regulatory authority with the mandate to regulate medical devices, if the legislation exists that empowers the regulatory authority, if there are established definitions for medical devices and IVDs, the existence of established principles of safety and performance, declaration of conformity by manufacturers, the basis for reliance and recognition, compliance with ISO 13485 requirements for medical devices, labelling requirements, advertising of medical devices, regulation of IVDs and post-market surveillance. The scale was Yes or No, with the possibility of explaining in detail where applicable. One staff member completed questionnaires for the three institutions (MCAZ, MLCScCZ and NMRL). Secondly, semi-structured interviews were conducted with key informants from the MCAZ, MLCScCZ and NMRL. Selected regulators from South Africa (SA), Tanzania and the WHO were selected to learn from them the best practices in their transition to regulate medical devices effectively. Semi-structured interview questions focused on the essential features of the WHO’s Global Model Regulatory Framework for Medical Devices (13).

Secondly, for the semi-structured interviews, open-ended ones were structured to explore the essential features of the medical devices regulatory system as recommended by the WHO Global Model Regulatory Framework and the Global Benchmarking Tool (GBT). Questions were structured to explore the existence and implementation of legal provisions and regulations for the following regulatory functions:

- regulatory system,
- market authorisation,
- licensing establishment,
- regulatory inspection (manufacturing site inspection),
- laboratory testing, and
- post-market surveillance.

To ensure the validity of the questions, the questions were adapted from the GBT. The GBT is the globally accepted standard for benchmarking regulatory authorities (14,15).

### Setting and Participants

The questionnaires were completed by representatives of the Medicines Control Authority of Zimbabwe (MCAZ), the Medical Laboratory and Clinical Scientists Council of Zimbabwe (MLCScCZ), and the National Microbiology Reference Laboratory (NMRL). The MCAZ is the NRA with the legal mandate to regulate medicines and allied substances. The MCAZ also regulates condoms and gloves as medical devices. The MLCScCZ is a health professional regulatory body that regulates the practices of medical laboratory professionals and some operations of medical laboratories. The MLCScCZ coordinates performance evaluations to grant IVDs access to national tenders related to HIV, TB, Malaria and STIs IVDs. The NMRL is the reference laboratory that conducts performance evaluations for market approvals that the MLCScCZ grants. Representatives regulating medical devices were purposively selected to complete the questionnaire to provide rich, relevant and diverse data pertinent to the research question (see file S1).

After developing a list of potential participants with input from key informants, regulatory documents and local knowledge, we used purposive sampling to identify eight participants from the MCAZ, MLCScCA, and NMRL to interview. We further supplemented this list by snowball sampling, reaching 12 interviewed participants—this brought the total number of participants from the MCAZ to four. Although the MCAZ has several units, the Medical Devices and Microbiology unit was selected as the interview focus because staff members in that unit are involved in market authorisation, manufacturing site inspections, laboratory performance evaluations and licensing activities. Staff members from the MLCScCZ (*N* = 4) were interviewed to gain an in-depth understanding of the IVD evaluation process. Staff members responsible for coordinating and performing laboratory testing of IVDs were interviewed at the NMRL (N=4)—table 1. The interviews were conducted face-to-face, and some were conducted remotely using Zoom and Microsoft Teams applications. All interviews were audio recorded after informed consent was obtained from participants. See Table 1 for participants’ characteristics, participants that completed the questionnaires and those that were interviewed. Regulators South Africa’s regulatory, South African Health Products Regulatory Authority (SAHPRA) and Tanzania, Tanzania Medicines and Medical Devices Authority (TMDA) were also interviewed to learn best practices and lessons learnt when the two countries established and implemented medical device regulations. The two countries were specifically chosen because of their tremendous medical device advancements in the Southern and Eastern African regions. Lastly, a WHO Regulatory Systems Strengthening representative was interviewed to understand the coverage of the WHO Global Model Regulatory Framework for Medical Devices in the Afro region and how it can be used to establish and strengthen medical device regulations. The idea was to use come up with a road map that is harmonised with other regional and international medical device regulations.

**Table 1:**
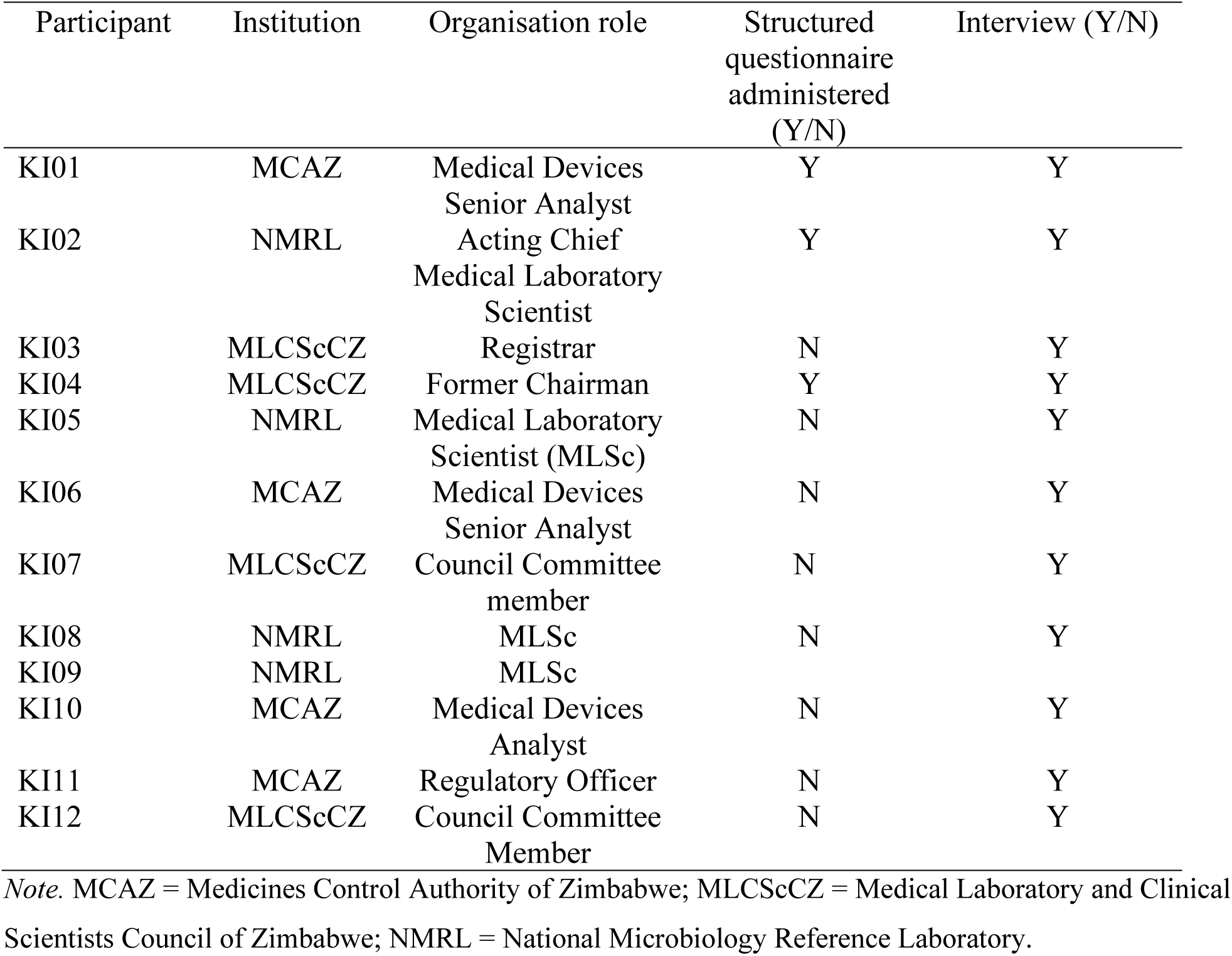
Characteristics of Interview Participants interviewed in Zimbabwe between June to November 2022

### Data Analysis

The Consolidated criteria for reporting qualitative research (COREQ) were used to ensure that all relevant information was included when preparing this manuscript. The criteria included the research team, study methods, context of the study, findings, analysis and interpretations (14). The researchers introduced themselves to the participants to avoid bias by stating their identities, credentials, occupation, gender, experience and training. The researchers did not have a relationship with any participants as an additional measure to mitigate bias. The study method was based on a content analysis theoretical framework to determine specific words, themes, or concepts within the qualitative data. Data were analysed using NVivo qualitative software version 12. Interview records were transcribed verbatim. Data was coded deductively using a coding frame and inductively by adding codes emerging from the data. CC and FB independently coded a subset of four transcripts each and then met to determine consensus and minimise intercoder variability for quality control purposes. The themes that emerged from the data analysis were acts, regulations and guidelines. For each theme, codes were developed for subthemes. The usefulness of these codes was tested by using this coding framework to code the first four transcripts in NVivo. Subsequently, the codes were discussed with and validated by a second researcher. Participants were assigned a code to maintain their anonymity, with codes reflecting their organisation (e.g., MCAZ KII7 was assigned to a participant from the MCAZ).

### Ethical Considerations

Ethical approval for this study was obtained from the Medical Research Council of Zimbabwe (MRCZ/A/2900). Participation was voluntary. Participants were able to stop the interview at any time without explanation. Written informed consent was obtained from each study participant before each interview. The interview content and the interviewee’s identity were kept anonymous.

## 3. Results

### 3.1 Section A: Questionnaire Findings

Based on questionnaire responses, it was found that at the time of this study, the MCAZ, MLCScCZ and NMRL did not have the explicit legal mandate to regulate medical devices, including IVDs.The MCAZ regulated condoms and gloves. The MLCScCZ, in collaboration with the NMRL, were conducting laboratory evaluations for market authorisation of IVDs for case management of HIV, Malaria, Tuberculosis, Syphilis, and Severe Acute Respiratory Syndrome Corona Virus 2 (SARS-CoV-2). Medical devices were not regulated, including IVDs for human and veterinary use, IVDs used in the public and private sectors, and IVDs for blood products. Participants from the MCAZ and MLCScCZ indicated that the regulations are still in the draft stage and need to be implemented. Regulatory functions focusing on premarket (assessment to grant market approval), marketing controls and post-market surveillance for medical devices (excluding condoms and gloves) and IVDs were not implemented.

### 3.2 : Results from Semi-structured Interviews

Stakeholders from Zimbabwe were interviewed to explore their perception of the current and future status of medical device regulation. Results from the interviews are presented in two sections. The first section includes interviews with Zimbabwe stakeholders, and the second includes interviews with regulators outside of Zimbabwe.

#### 3.2.1. Results from Interviews with Stakeholders from Zimbabwe

Semi-structured interviews were conducted with 15 participants, 12 of whom were critical stakeholders in medical device regulation in Zimbabwe. The themes identified from the interviews were categorised under critical components of a regulatory system. Transcripts were reviewed first, and data were organised by categories that had emerged during the review, i.e., connected verbatim quotes were categorised. We then reviewed and interpreted the quotes within each category. This enabled us to identify six main themes across our interview data. The six themes included: limited legal framework; limited regulatory capacity; conformity assessment not proportionate to risk classification, lack of post-market surveillance; the need to improve the medical devices regulatory system; and collaboration of regulatory authorities. See Table 2 for themes and quotes from participants capturing the themes.

**Table 2:**
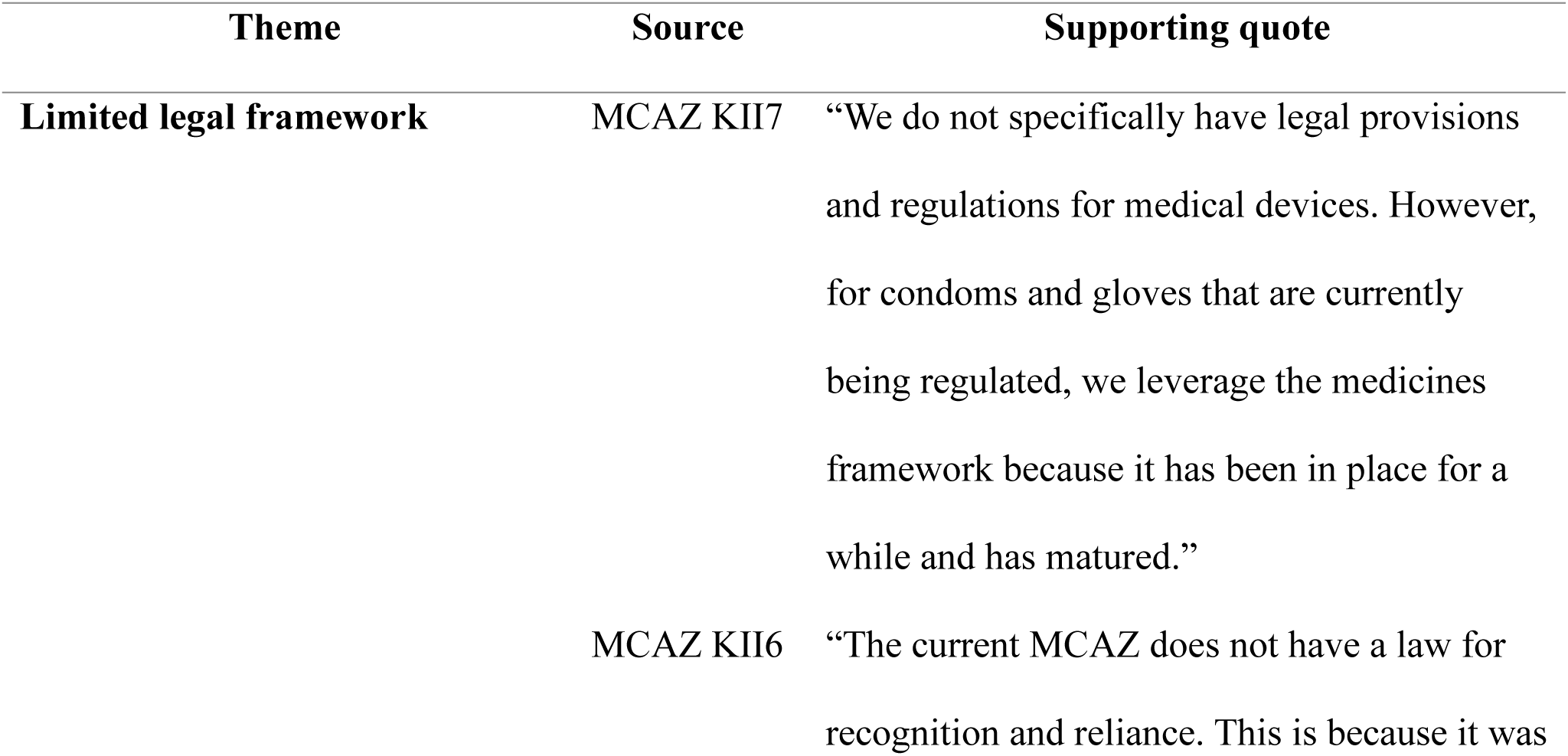
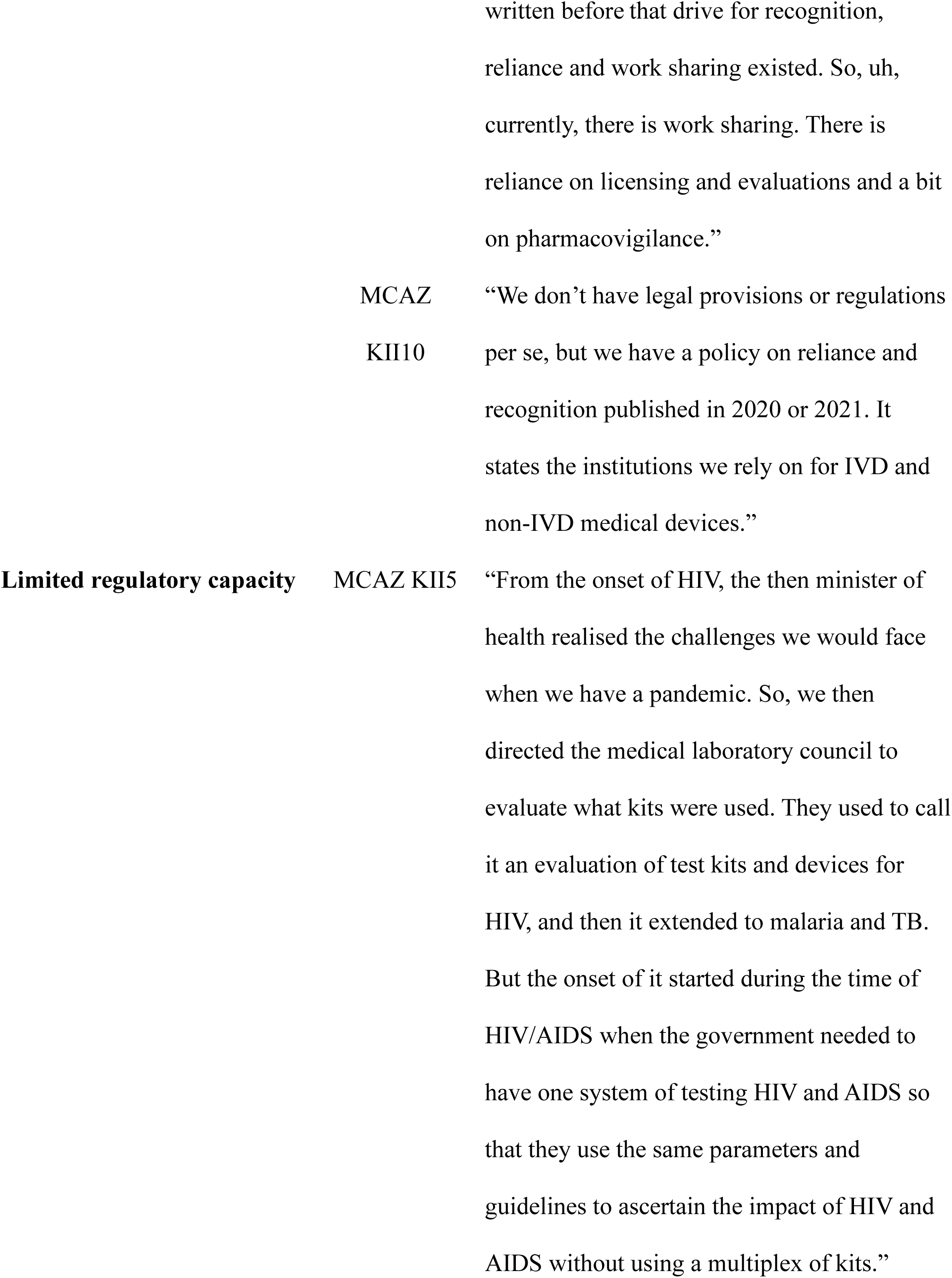
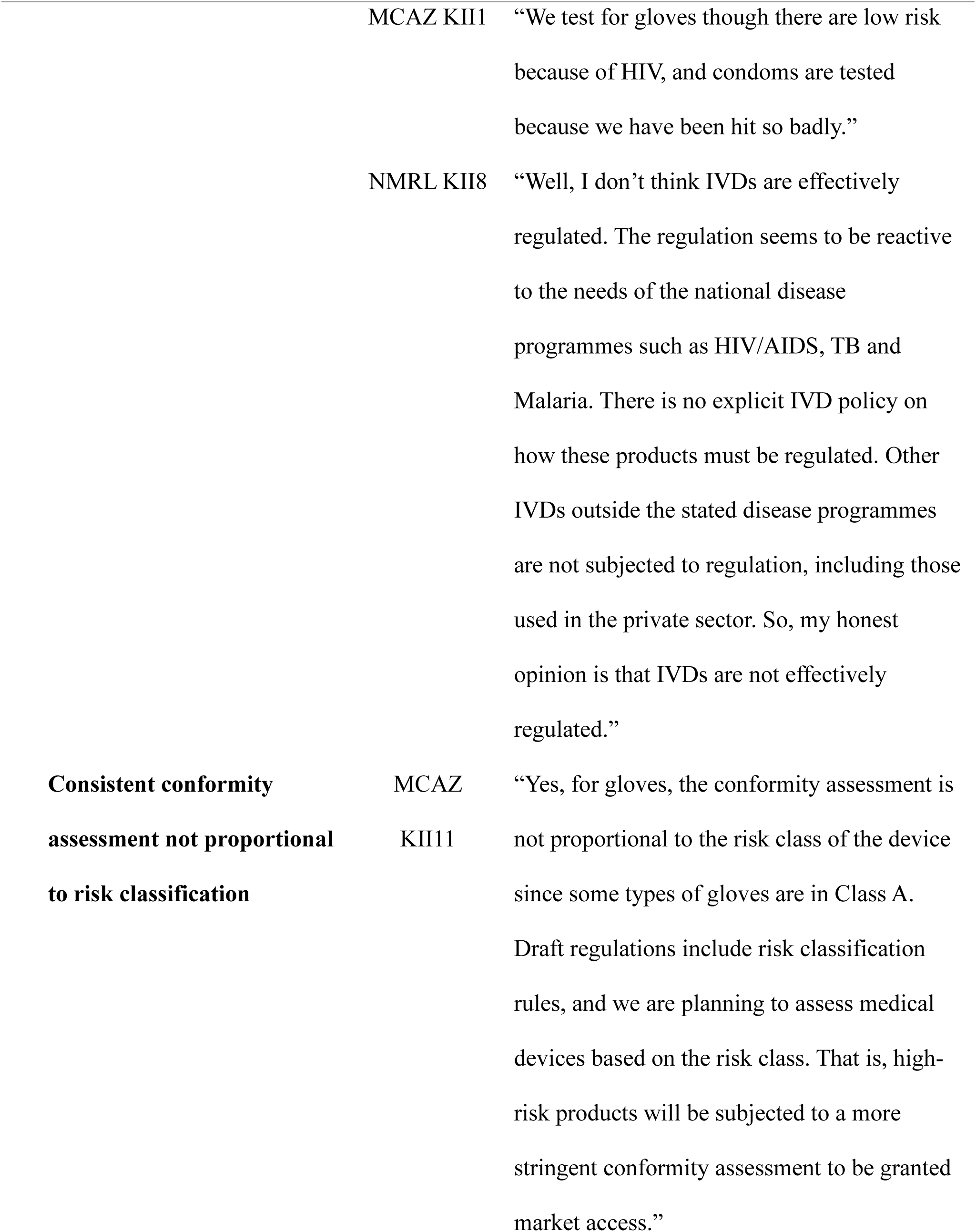
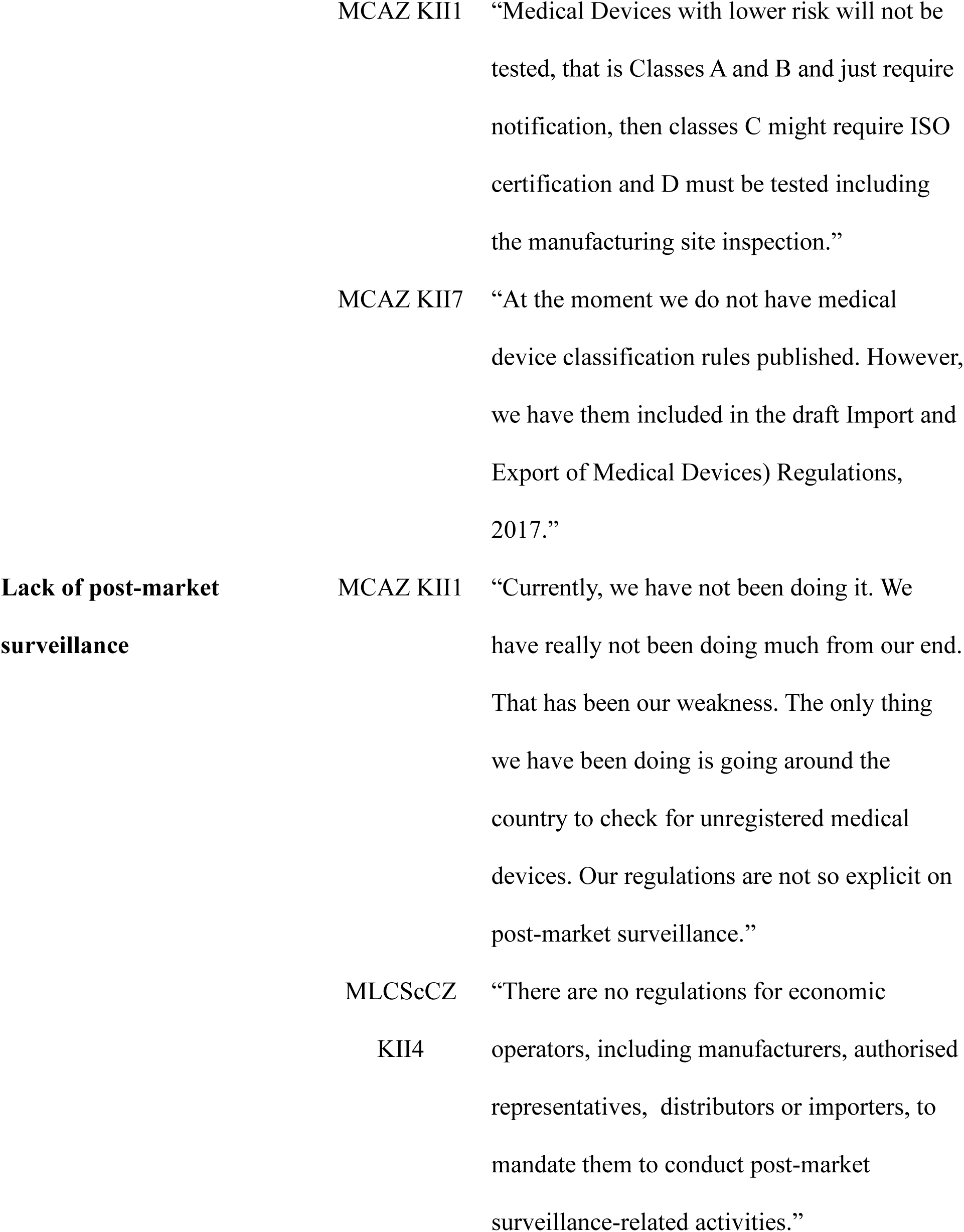
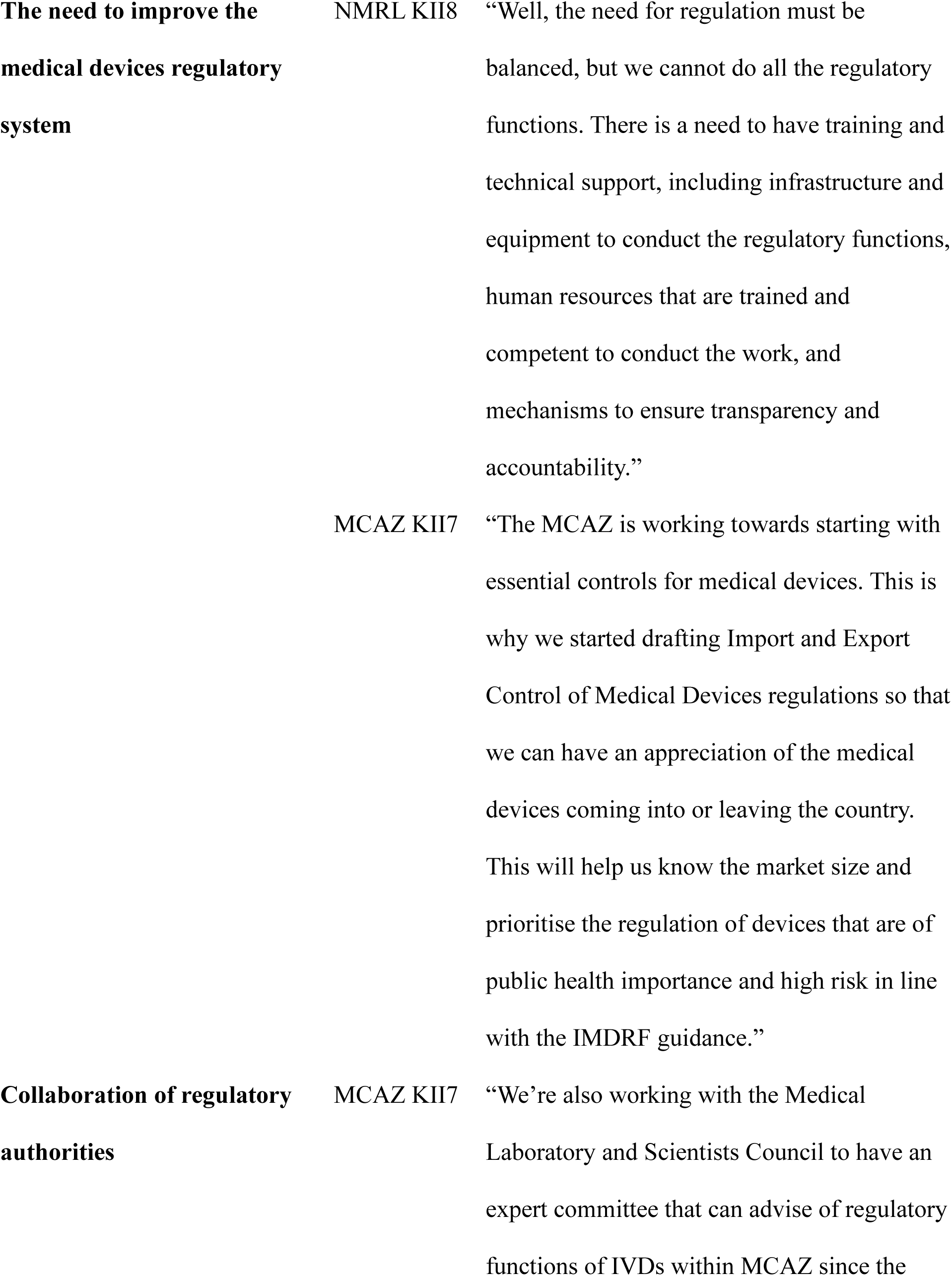
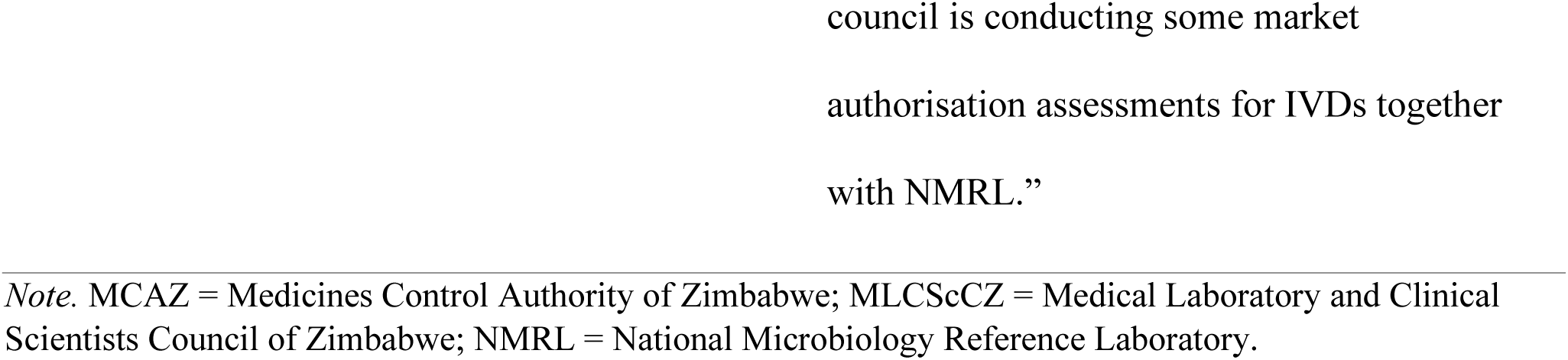
Main themes emerging from the interviews of medical device regulatory stakeholders in Zimbabwe conducted between June to November 2022

##### 3.2.1.1 Theme 1: Limited legal framework

The regulation of medical devices in Zimbabwe is based on something other than an effective and efficient legal and policy foundation. The MCAZ assumed the mandate to regulate medical devices, including IVDs using the Medicines and Allied Substance Control Act (MASCA). On the other hand, the MLCScCZ assumed to mandate to regulate IVD medical devices based on the Health Professions Act. The MASCA was initially gazetted following the medicines framework and missed some critical medical device regulation aspects. For example, the law must clearly state the scope of regulated medical products and identify the entities subject to regulation.

Additionally, there is no general requirement that only medical devices that are safe, of acceptable quality, and performance must be authorised to access the Zimbabwean market. However, during the interviews, MCAZ respondents noted that the MCAZ regulated condoms and gloves as part of medical devices, stating: “*We do not specifically have legal provisions and regulations for medical devices. However, for condoms and gloves that are currently being regulated, we leverage the medicines framework because it has been in place for a while and has matured” (MCAZ KII7).* The MLCScCZ was conducting registration for IVDs specific for priority diseases for the national tendering process, *“But not to say they have the powers to do it, the council would report from the of the approved products for tendering process with data produced from the evaluation by the experts’ committee”* (MLCScCZ KII4).

The MCAZ had drafted regulations for the Import and Export of Medical Devices and In Vitro Diagnostic Medical Devices, but these were yet to be approved during the interviews. The MCAZ had plans to amend the MASCA to include all health products that require regulation: “*Exactly. Currently, there is a bill that has been drafted, so you will see MCAZ changing its name to a name that encompasses other products. Currently, the name implies control of medicines only, but there are discussions with the Ministry of Health for the name to encompass all medical products.”* (MCAZ KII1). It was unclear if the NMRL had a legal mandate to conduct laboratory testing for regulatory purposes: *“Ummm, it is unclear as far as legislation is concerned. However, it has been known within NMRL and the Ministry of Health that the NMRL conducts performance evaluations for IVDs for the Medical Laboratory and Clinical Scientists Council of Zimbabwe to register products procured through national tendering processes”(NMRL KII8).* Furthermore, the legislation that mandates NMRL to perform laboratory testing functions for regulatory purposes was unclear. One interviewee responded to the question of the existence of the Act of Parliament that established the NMRL if it included conducting regulatory functions for IVDs by saying, *“ I have heard there is an Act Parliament, but we have failed to locate it. So, it is unclear whether the act exists and what mandates are given to NMRL according to the act.” (NMRL KII8)*.

Therefore, the legislation in Zimbabwe needs to be more explicit on the institutions involved in medical device regulation and that medical devices, including IVDs, that meet specific safety, quality, and performance requirements must be authorised to access the market.

##### 3.2.1.2 Theme 2: Limited regulatory capacity

The MCAZ was regulating condoms and gloves only at the time of the interview. Other medical devices were not regulated. The capacity to regulate IVD medical devices in the MCAZ seems limited. They were engaging the MLCScCZ to have an experts committee to assist with the regulatory functions of IVDs within the MCAZ in the future: “*The technical committee approves, and then they are shared with external stakeholders, at some stage, they should be a stakeholder consultation that includes MLCScCZ, HPA, Ministry” (MCAZ KII1).* The MLCScCZ also showed its challenges in regulating IVDs and was keen to work with the MCAZ since it has some expertise in other health products such as medicines, condoms and gloves: “*So we’re going to be looking at a manufacturing site, which manufacturer bringing in IVDs. I think we’ll do it like what MCAZ does for drugs cause they do a thorough job cause I worked with those guys. They do go even to where those things are being manufactured to see what they’re saying is what they supposed to do”* (MLCScCZ KII3). During the interviews, respondents noted that the MLCScCZ used only performance verification results from the NMRL to make registration decisions for IVD registration. The NMRL had challenges related to staff not being trained on the regulatory functions of the laboratory, non-availability of Clinical Laboratory and Standards Institute guidelines and testing panels (reference samples), and testing protocols were not standardised to meet the CLSI guidelines: “*Yes, because we do not have well-characterised specimen panels, and we have to rely on results from other institutions, such as condemned blood from the National Blood Services.”* (NMRL KII9).

Overall, the institutions currently regulating medical devices have limited capacity in terms of resources, expertise and processes to regulate medical devices, including IVDs, effectively.

#### 3.2.3. Theme 3: Conformity Assessment not proportional to risk classification

The MCAZ conducted conformity assessments for condoms and gloves. However, there are no criteria for the classification of medical devices using established classification rules to guide the level of scrutiny the product must be subjected to. *“At the moment we do not have medical device classification rules published. However, we have them included in the draft Import and Export of Medical Devices Regulations, 2017”* (MCAZ KII7). However, some gloves are Class A and are regulated with the same stringency as Class C devices: *“Yes, for gloves, the conformity assessment is not proportional to the risk class of the device since some types of gloves are in Class A. Draft regulations include risk classification rules, and we are planning to assess medical devices based on the risk class. That is, high-risk products will be subjected to a more stringent conformity assessment to be granted market access.”* (MCAZ KII11).

No risk-based approach is currently in place to classify medical devices that work as a precursor to deciding on the level of scrutiny the medical devices will be subjected to during conformity assessments.

#### 3.2.4. Theme 4: Lack of post-market surveillance

The MCAZ was not conducting formal PMS for gloves and condoms besides routine inspections following the pharmacovigilance framework of medicines. *“Currently, we have not been doing it. We have really not been doing much from our end. That has been our weakness. The only thing we have been doing is going around the country to check for unregistered medical devices. Our regulations are not so explicit on post-market surveillance ….” (MCAZ KII1).* The MLCScCZ gets reports on IVDs, but the process is poorly documented. There are no documented obligations for economic operators (manufacturers, importers, authorised representatives, distributors and wholesalers) to conduct PMS activities: *“Yes, from the regulatory side and any economic operator looking at the manufacturers, authorised representatives, distributors or in importers, such things are not clearly defined on how the process should go about”* (MLCScZ KII03).

PMS regulations and procedures are not documented. Therefore, there is no assurance that the products on the market and in use are still safe, of acceptable quality and performance.

#### 3.2.5. Theme 5: The need to improve the medical devices regulatory system

There was a consensus from the participants that medical device regulation needed to be improved. Regulation of medical devices has been suboptimal: “*Well, I don’t think IVDs are effectively regulated. The regulation seems to be reactive to the needs of the national disease programmes such as HIV/AIDS, TB and Malaria. There is no explicit IVD policy on how these products must be regulated. Other IVDs outside the stated disease programmes are not subjected to regulation, including those used in the private sector. So, my honest opinion is that IVDs are not effectively regulated”* (NMRL KII8). The MCAZ has drafted regulations and is amending the MASCA: “*The MCAZ is working towards starting with essential controls for medical devices. This is why we started drafting Import and Export Control of Medical Devices regulations so that we can have an appreciation of the medical devices coming into or leaving the country. This will help us know the market size and prioritise the regulation of devices that are of public health importance and high risk in line with the IMDRF guidance” (MCAZ KII7). My desire is to see the regulation of IVDs harmonised with international best practices. For example, we can use best practices such as those applied for WHO Prequalification and other mature regulatory authorities* (MLCScCZ KII11). There is a general willingness from stakeholders to improve the regulation of medical devices, and suggestions have been given on this can be done.

#### 3.2.6. Collaboration of regulatory authorities

Despite unclear legislation on the institutions with the mandate to regulate medical devices, efforts have been made to allow for collaboration between the MCAZ and MLCScCZ. The two institutions are finding each other, recognising the expertise and role each institution can contribute to effectively regulating medical devices. *“We’re also working with the Medical Laboratory and Scientists Council to have an expert committee that can advise on regulatory functions of IVDs within MCAZ since the council is conducting some market authorisation assessments for IVDs together with NMRL”* (MCAZ KII7). The role the NMRL plays in conducting laboratory testing for regulatory purposes is also recognised by the MCAZ: “… *the NMRL will continue doing what they were doing. But they’ll work more like, um, assessment bodies where they will test on behalf of the regulator. Then they give The results to the regulator so that the regulator only does the evaluation process, but not testing of the, of the kits.”* (MCAZ KII6). Another respondent had this to say, “ *This requires a pragmatic approach, that includes building the capacity of regulators, infrastructure, a collaboration between institutions based on their expertise, and leadership. We need now to think of this as a system, not as a way of protecting specific professions but as putting the safety of the patients at the centre* (MLCScCZ KII11)”. The responses clearly show the need to collaborate and the willingness of key stakeholders to come together to establish and strengthen medical device regulations.

#### 3.2.2 Results from Interviews with Regulators Outside of Zimbabwe

One regulator from SAHPRA and the other from TMDA were interviewed to understand the journey SAHPRA and TMDA went through when they transitioned from regulating medicines to including medical devices in the scope of regulated medical products. Lessons learnt and best practices were documented to aid in drafting the roadmap for developing medical device regulations in Zimbabwe and for harmonisation purposes. The WHO representative was interviewed to understand the implementation and coverage of the Global Model Regulatory Framework for Medical Devices and how countries can use it to strengthen medical device regulation. Tables 3 and 4 show the characteristics of the interviewees and the themes that emerged from the interviews.

**Table 3:**
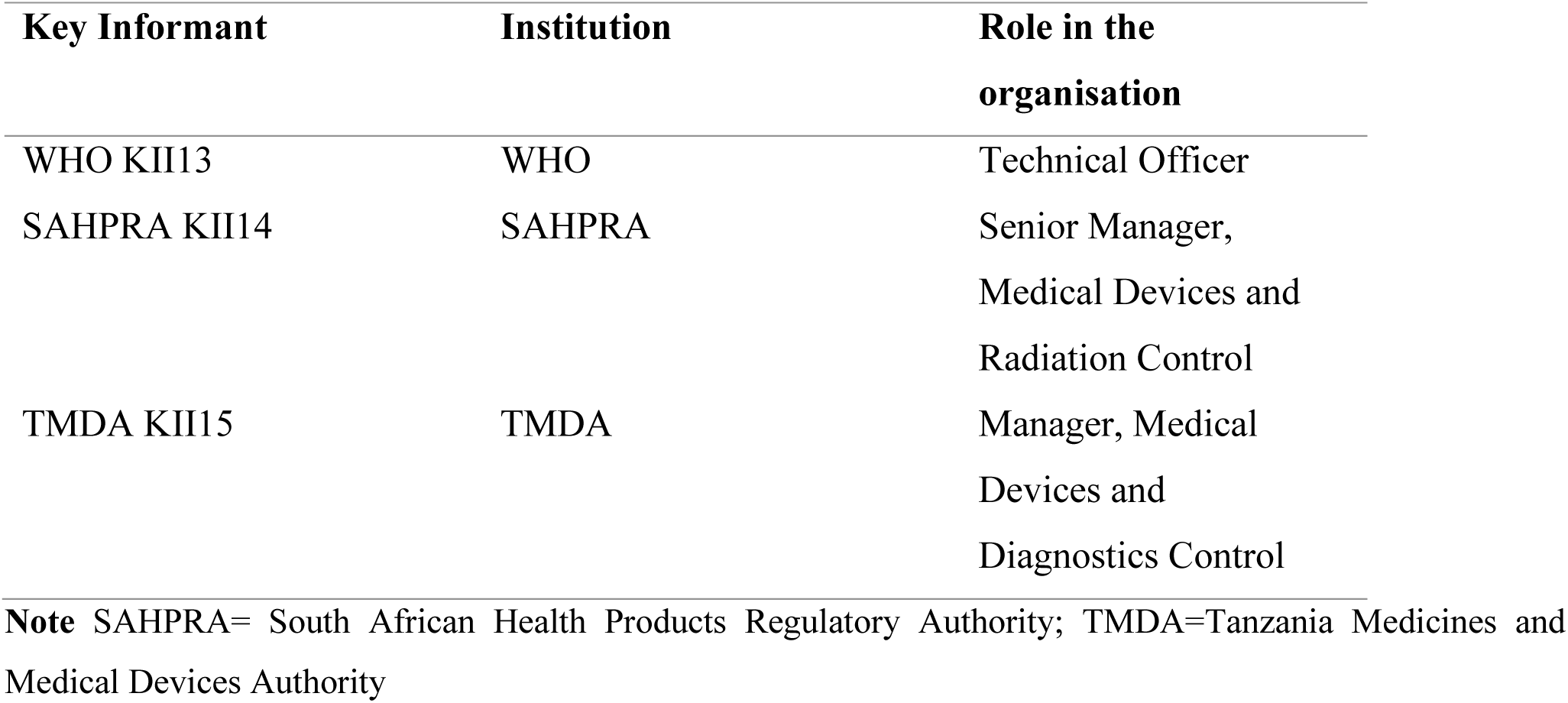
Characteristics of Key Informants that Participated in Interviews from outside of Zimbabwe between June and November 2022

| Key Informant | Institution | Role in the organisation |
| --- | --- | --- |
| WHO KII13 | WHO | Technical Officer |
| SAHPRA KII14 | SAHPRA | Senior Manager, Medical Devices and Radiation Control |
| TMDA KII15 | TMDA | Manager, Medical Devices and Diagnostics Control |

**Note** SAHPRA= South African Health Products Regulatory Authority; TMDA=Tanzania Medicines and Medical Devices Authority

**Table 4:**
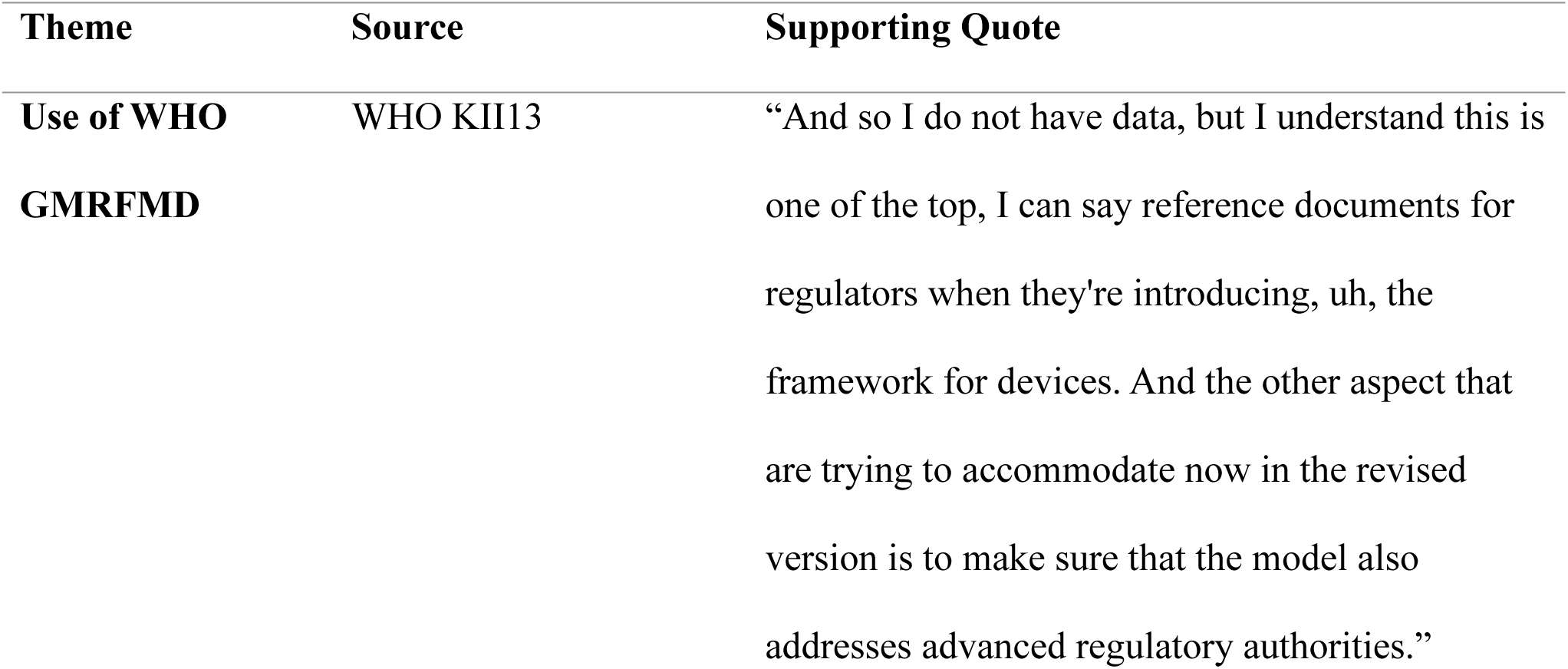
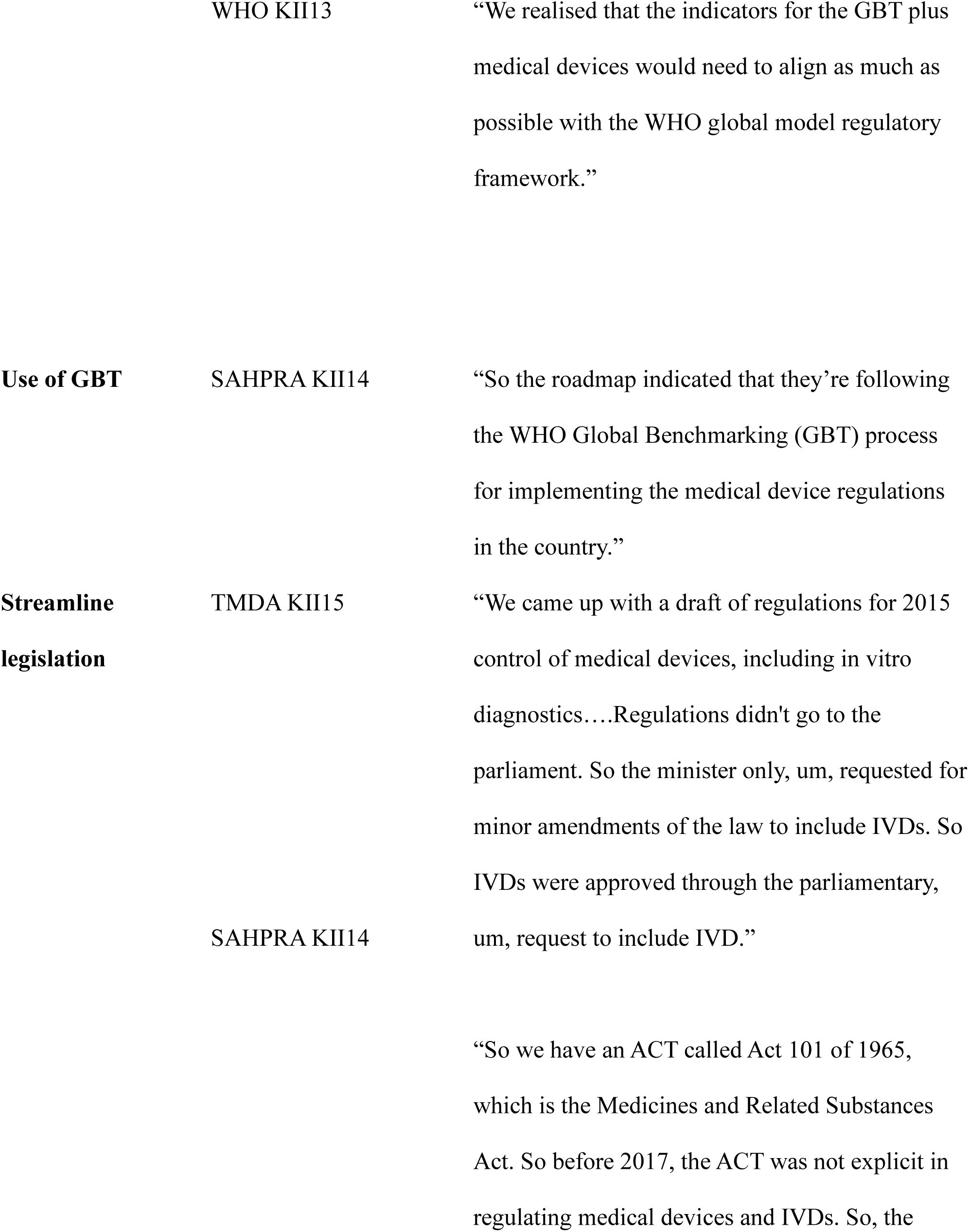
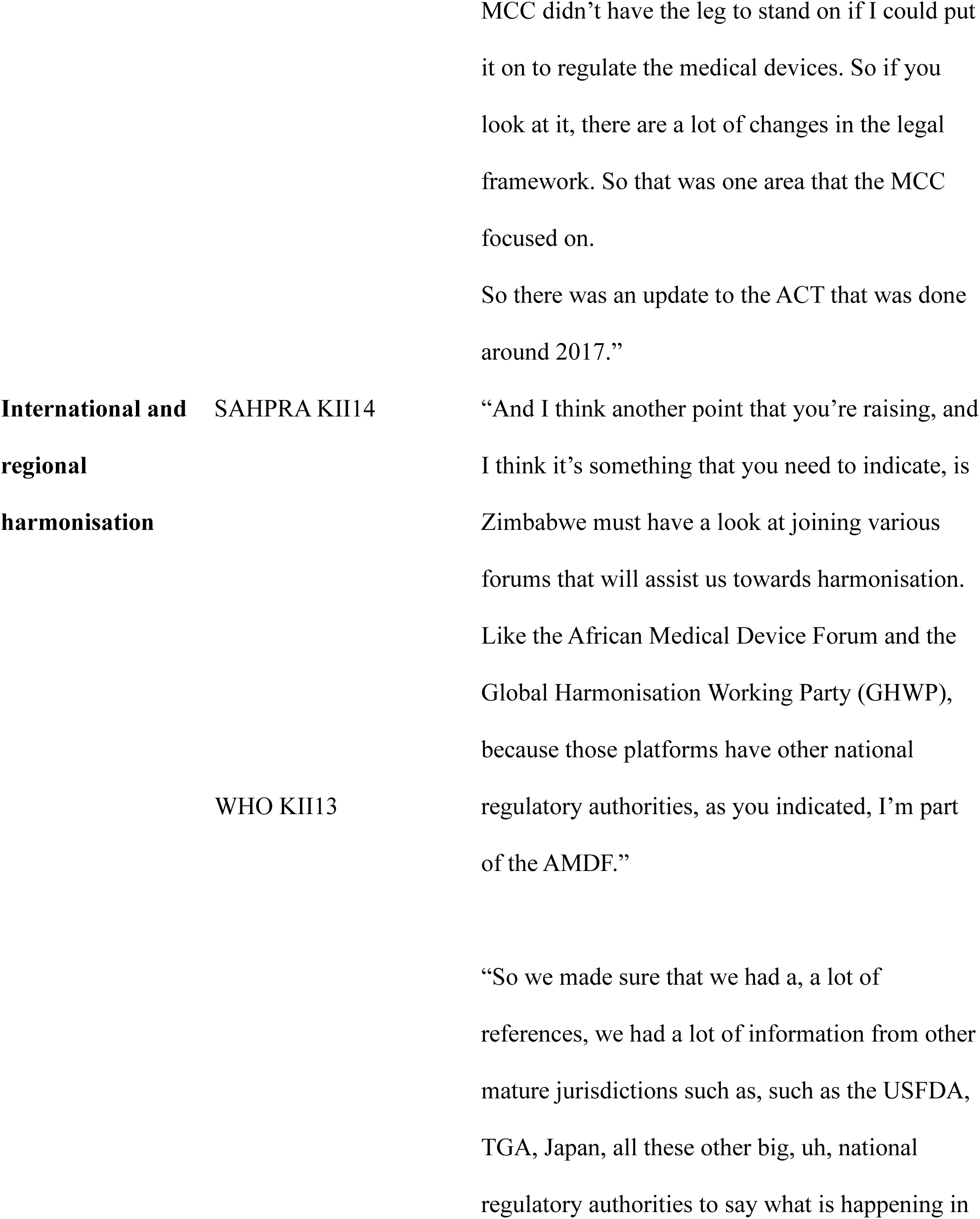
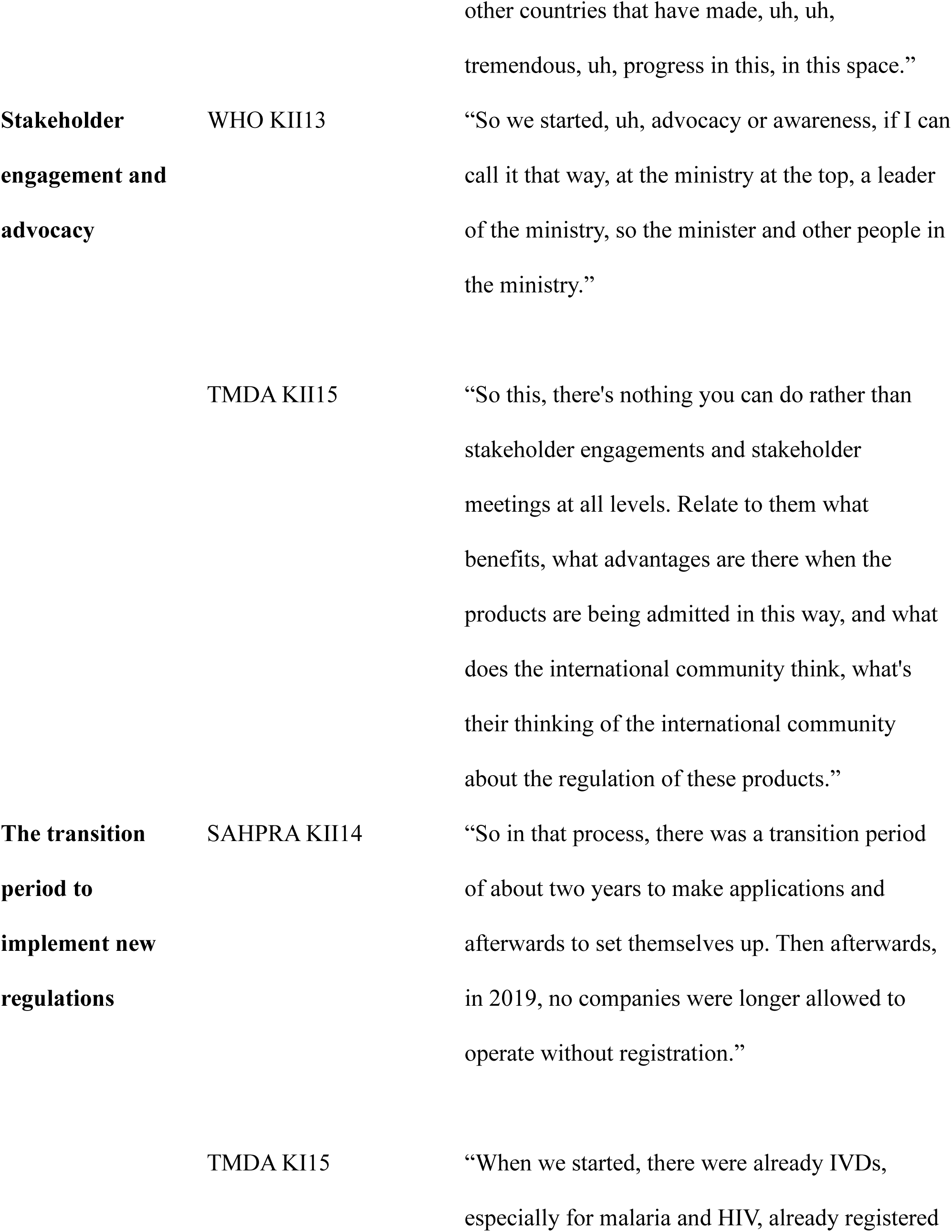
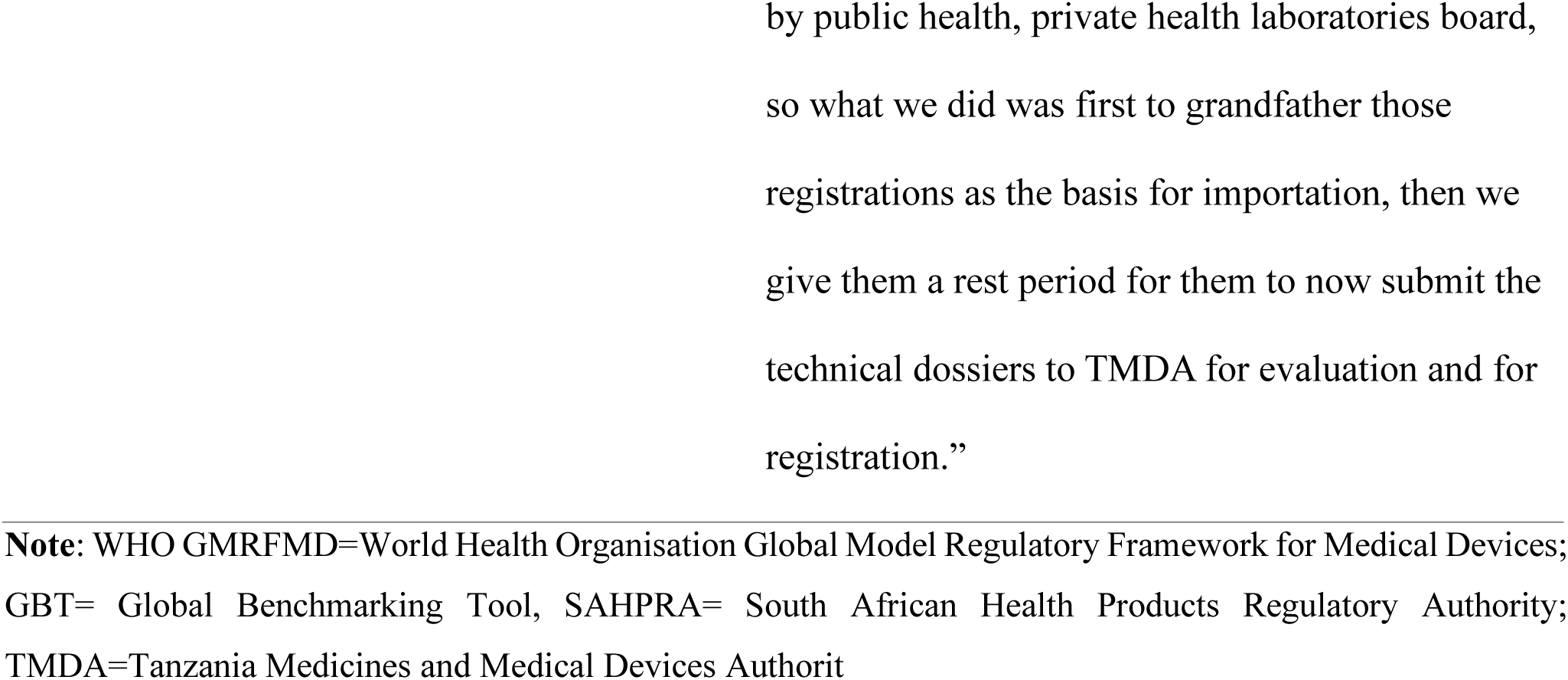
Main themes emerging from the June to November 2022 interviews on Key Informants from outside Zimbabwe.

##### 3.2.2.1 Use of WHO GMRFMD

The WHO GMRFMD was recognised as a reference to establish and improve medical device regulations. In a survey that the WHO conducted in the Afro region, about 60% of the countries had awareness and used the framework: “ *So we are looking at maybe 58 % understanding of the model. And also, uh, using it to, as a guidance, uh, as a reference document, uh, for the regulation of medical devices and IVDs. It’s unfortunate that we didn’t conduct a global survey.”* (WHO KII13). The WHO GMRFMD is an acceptable framework that countries can use to establish and strengthen medical device regulations.

##### 3.2.2.2 Use of GBT

South Africa used the GBT to map its way to improving the medical devices regulatory system. The GBT has indicators that are used to benchmark the regulatory system and the development of an institutional development plan that can be implemented and monitored: *“ So the roadmap indicated that they’re following the WHO Global Benchmarking (GBT) process for implementing the medical device regulations in the country (*SAHPRA KII13)”. Revision of the GBT to include medical devices has presented an opportunity for the WHO GMRFMD to be revised to cover both basic and advanced controls for medical devices. The WHO representative said, *“ And, so the discussion in the GBT indicators triggered revision. And this is why there is an ongoing discussion to update and revise the model* (WHO KII13)”. The WHO GMRFMD needs to be utilised together with the GBT as the two work together for institutions to develop and strengthen their regulatory systems.

##### 3.2.2.3 Streamline legislation

NRAs from South Africa and Tanzania had to amend their legal framework to ensure medical device regulation was above board. In the South African case, the law was amended to allow the name change of the NRA and the law to regulate medical devices. “ *So while it was still MCC before 2017, some regulations were published, and then the industry was pushing back on them and an update to the legislative framework, which is the ACT. So we have an ACT called Act 101 of 1965, which is the Medicines and Related Substances Act. So before 2017, the ACT was not explicit in regulating medical devices and IVDs. So, the MCC didn’t have the leg to stand on if I could put it on to regulate the medical devices.” (SAHPRA KII13).* The Tanzanian NRA representative had this to say about the change in the legal framework, *“ So the minister only, um, requested for minor amendments of the law to include IVDs. So IVDs were approved, uh, through the parliamentary, um, request to include IVD. So that was included, amendment of the definition, the inclusion of the risk classes of devices. And then, uh, it gave the minister also power to make other regulations. So we used that clause to develop specific regulations for medical devices. And so, uh, the minister signed the regulations in 2015(TMDA KII15).”*The legislation is the foundation for regulation to be established and enforceable.

##### 3.2.2.4 International and regional harmonisation

Tanzania conducted an assessment to devise ways of harmonising the medical device regulations. The assessment was conducted by looking at regulations from mature regulatory authorities such as the United States of America Food and Drug Administration (USFDA), the Australian Therapeutic Goods Administration (TGA), and the Japanese Pharmaceuticals and Medical Devices Agency (PMDA). “ S*o we made sure that we had a, a lot of references, we had a lot of information from other mature jurisdictions such as, such as the USFDA, TGA, Japan, all these other big, uh, national regulatory authorities to say what is happening in other countries that have made, uh, uh, tremendous, uh, progress in this, in this space for harmonisation purposes (WHO KII15).*” Tanzania’s regulatory authority framed its regulations following the IMDRF framework to ensure harmonisation. *“ Of course, it was very much mirrored from medicine’s regulatory framework, as well as at that time, the IMDRF mode of regulation to ensure harmonisation.”* (WHO KII15). SAHPRA representative encouraged Zimbabwe to join harmonisation initiatives to learn best practices from other regulatory authorities. “ *And I think another point that you’re raising, and I think it’s something that you need to indicate, is Zimbabwe must have a look at joining various forums that will assist us towards harmonisation. Like the African Medical Device Forum and the Global Harmonisation Working Party (GHWP), because those platforms have other national regulatory authorities, as you indicated based, I mean, I’m part of the AMDF. So there is already another regulator in that platform* (SAHPRA KII13).” South Africa and Tanzania harmonised their regulations with other stringent regulatory authorities, which is commendable.

##### 3.2.2.5 Stakeholder Engagement and Advocacy

Stakeholder engagement and advocacy were conducted during the medical device regulations formulation. The engagement was done at different levels, from technical departments to policymakers in the Tanzanian context. “*So if the minister is on our side, if the policymakers, if the, uh, permanent secretary of the Minister of Health and the directors at the Minister of Health understand where we are going wrong, then definitely we would have supposed to say, you know, guys, we need to correct this. So we started, uh, advocacy or awareness, if I can call it that way, at the ministry at the top, a leader of the ministry, so the minister and other people in the ministry.”* (WHO KII13). In the South African context, the mandates were clearly defined based on legislation, so engagement with other stakeholders in regulating medical devices was not complicated. “ *Cause that’s not their mandate. So exactly. Now, for example, our act has radioactive medical devices. We also have national regulatory authority. But they cannot take the health product all these years; they were just sitting in the Ministry of Health. They couldn’t take it. They can look after the isotopes and this big Eskom stuff that produces radiation, but they couldn’t come across that boundary to come and do medical devices and IVDs because the legal framework prohibits them to do so* (SAHPRA KII14)”. It is clear from the interviews that stakeholder engagement and advocacy are essential so that parties involved in medical device regulations know their mandates and that the leadership is involved in the process.

##### 3.2.2.6 Transition period to implement new regulations

When new regulations were promulgated in South Africa and Tanzania, there was a transition period for economic operators to comply with the new regulations. This was done so that there was no disruption in access to medical devices. *“ So in that process, there was a transition period of about two years to make applications and afterwards to set themselves up. Then afterwards, in 2019, no companies were longer allowed to operate without registration, so what we did is that company will send applications cause the applications and the guidelines were drafted, companies will send applications, and then SAHPRA would give them a letter of acknowledgement so that there is no disruption in the business because there were lots of applications that came about 600, 700 in a year.”* (SAHPRA KII14). Tanzania allowed IVDs registered by the Private Health Laboratory Board (PHLB) to access the market and gave a transition period for the manufacturers to formalise the registration of their IVDs. *“ So we had to rely on that status, at least for the first year, before we can allow them to apply for full registration. And also, even when we were grandfathered in there when we were now registering them, we used to recognise the performance that was being carried out by the PHLB because what PHLB, so their mode of regulation was only on performance, while ours is around all the areas of regulation, including site performance.* (TMDA KII15).” The introduction of regulations mustn’t work as a barrier for the population to access medical devices when new regulations are introduced.

## Discussion

Medical device regulation ensures that medical devices approved for use meet the specified safety, quality and performance requirements to protect public health (7). Regulation must be based on a solid legal framework to be enforceable. The legislation details the mandate of the national regulatory authority, regulatory system, scope of regulated medical products, definitions of regulated medical devices, and regulatory functions of the NRA or other institutions. Regulations interpret the legislation, law or act and allow the NRA to establish and implement principles to ensure the safety and performance of medical devices and evaluate if the medical devices used in public and private sectors are regulated (2). One of the significant findings of this study was that Zimbabwe does not have a formal medical devices regulatory system. Medical devices other than condoms and gloves, donated medical devices, IVD medical devices for human and veterinary and human blood products are not regulated. Hubner et al. had a similar finding when they investigated the landscape of medical device regulatory systems in East, Central and Southern Africa. However, Hubner et al. concluded that Zimbabwe had a legal framework for medical devices (9). This contradicts our finding, as the participants stated that the law is not yet amended to regulate medical devices. The differences may be due to methodological differences, as Hubner et al. used a literature review. Considering the shortage of literature in the Zimbabwean context, the granularity of information is limited. The participant who completed the questionnaire from MCAZ responded to question number two on the questionnaire, *"Does your institution has a legal mandate to regulate medical devices, including IVDs?"* as *"Not yet. The regulations are still in draft stage and not yet approved/gazetted for implementation"*(MCAZ KII1).

Additionally, the legal framework (MASCA) was made specifically for medicines and allied substances but did not have a formal definition of a medical device. The WHO recommends it in the GMRFMD for the legal framework to clearly define medical devices and the scope of these devices that must be regulated to ensure that only medical devices that meet conformity assessment are granted market authorisation and approved for use (13). The difference between Hubner et al.’s and our study on the presence of the legal framework may be due to the way questions were structured to collect this information. In our study, semi-structured interviews complemented questionnaires to triangulate the findings from questionnaires and semi-structured interviews. Additionally, we utilised a case study strategy to probe deeply and analyse the myriad phenomena that constitute the complexities of the medical devices regulatory system. Hubner et al. used a literature review methodology. Although there was a contrast, some similarities were observed in these two studies. For instance, both researchers identified the following deficiencies: non-existence of the definition of medical device, risk-based classification of medical devices, and essential principles. Essential principles, conformity assessment, and registration components were found to be in place for gloves and condoms for both studies (9).

The MCAZ had drafted regulations for the Import and Export and Export of Medical Devices and In Vitro Diagnostic Medical devices, but they were yet to be approved. It is unclear when these regulations will be approved: “ *As for the timelines, we have played our part. We have drafted, and the approval is with the ministry; that’s where delays are. Our team has been pursuing, still pursuing. The ones that I am certain are going to be gazetted but outside of the scope of the study are for the regulation of blood and blood products.The ones for medical devices and IVDs I don’t know..*” (MCAZ KII1). “*In terms of, uh, the timeline, uh, in terms of MCAZ as a timeline, if we, if we had entirely on us, probably would’ve been like December. But now there’s so many things at play”* (MCAZ KII6). The delays in approving the draft regulations may be due to a lack of autonomy by the MCAZ to make approval decisions. On the other hand, these draft regulations are not supported by a sound basis in law that must include definitions of the products within its scope and identify the entities subject to regulation (13).

This finding implies that medical devices are not effectively regulated except for condoms and gloves; medical devices, including IVD medical devices used in the private and public sector, donated from donors, stains for microscopy and medical devices for veterinary use are not regulated. IVDs for detecting and monitoring HIV, Malaria, TB, Syphilis and COVID-19 are somehow regulated but ineffectively. IVDs are not effectively regulated because the conformity assessment ensures that the products are safe and of acceptable quality, and performance is not standardised. Only performance characteristics were verified. There was no technical documentation assessment, manufacturing site inspections or labelling review to complement performance evaluation. Therefore, there is no assurance that medical devices that are safe, of acceptable quality and perform as intended are marketed and used for the Zimbabwean population.

To understand the context of how the MLCScCZ ended up conducting some IVD regulations, it was indicated that MLCScCZ assumed the role of responding to the need for procurement requirements to ensure that IVDs had to go through an evaluation for them to be eligible to participate in national tendering processes for HIV IVDs. The MLCScCZ was granted the authority by the health minister to coordinate IVD registration for public national tendering processes to facilitate procurement. In response to the HIV pandemic, the minister of health realised the challenges faced with HIV testing due to non-regulated HIV test kits. The minister directed the MLCScCZ to coordinate the evaluation of HIV test kits for national tendering processes and the selection of test kits for use as part of the HIV Testing algorithm. These developments culminated in the establishment of the NMRL at Harare Hospital as it was deemed to have the expertise to conduct the evaluations to verify the performance of priority IVDs. Respondent KI04 said, “*From the onset of HIV, the then minister of health realised the challenges we would face when we have a pandemic. So, we then directed the medical laboratory council to evaluate what kits were used. They used to call it an evaluation of test kits and devices for HIV, and then it extended to malaria and TB. But the onset of it started during the time of HIV/AIDS when the government needed to have one system of testing HIV and AIDS so that they use the same parameters and guidelines to ascertain the impact of HIV and AIDS without using a multiplex of kits”* (MLCScCZ KII4).

On the other hand, the MCAZ started regulating gloves and condoms as IVDs in response to the HIV pandemic despite not having the legal mandate. Respondent MCAZ KII1 highlighted this point by saying, "*We test for gloves though there is low risk because of HIV. Condoms are tested because we have been hit so badly"*.

The MLCScCZ appreciated that they did not have the mandate to regulate IVDs but "claimed" it was MCAZ’s mandate, but the MCAZ was not regulating the IVDs. Therefore, there was a vacuum., *"I think things might improve because it’s like so, whether we like it or not, MCAZ does have the regulatory powers to do things we’re talking about, and then we try to do it as far as soon as possible." (MLCScCZ KII3).* A single coordinated approach that establishes roles and responsibilities makes formulating and enforcing regulations more feasible. The acknowledgement of MLCScCZ that it does not have the legal mandate to regulate IVDs shows some progress from when a similar study was conducted by Dancombe et al. The study investigated the stakeholder perceptions on current and future regulation of HIV Self-Testing IVDs in Malawi, Zambia and Zimbabwe. It was concluded that Zimbabwe did not have clarity on its mandate to regulate the HIV Self Testing IVDs; MCAZ and MLCScCZ both claimed the mandate. In the same study, one key informant from the MCAZ felt the need for HIV Self Test IVDs to be regulated by only one regulator and that there must be some collaboration with the MLCScCZ. There is an expectation that the MLCsCZ will work with MCAZ to regulate IVDs through an advisory committee from the MLCScCZ that will advise on IVD regulatory functions. Respondent MLCScCZ KI03 stated, "*But I think I’m happy to advise that we have now agreed that the council was trying to collaborate with MCAZ. Terms of reference for council members to be part of the advisory experts’ committee within MCAZ are in their draft format."*

The realisation by the MLCScCZ of the need to collaborate with the MCAZ on IVD medical device regulation is a step in the right direction to strengthen collaboration and coordination of IVD medical device regulation. The representative from NMRL indicated that it needed to be clarified if NMRL had the legal mandate to perform laboratory testing to support market authorisation of IVDs. Legal provisions must be implemented to ensure that the NMRL can formally conduct laboratory testing for IVDs.

Conformity assessment is conducted before and after a medical device is placed on the market. Post-market surveillance of devices in actual use ensures that the medical device continues to meet the safety and performance requirements and that the benefit-risk ratio maintains public confidence (5). The conformity assessment must be proportional to the risk classification of the medical device. The MCAZ assessed the dossiers to verify if the manufacturer’s claims about the product were substantiated by analytical and clinical evidence where applicable, manufacturing site inspection, and product performance evaluation for gloves and condoms. “ *The assessment is conducted if the product meets the requirements, the recommendation is made to the registration committee for decision-making for granting market authorisation. The samples are tested, and if they meet the performance characteristics, a provisional approval is granted, pending manufacturing site inspection”* (MCAZ KII1). It is unclear why MCAZ prioritised the regulation of gloves considering that there are classified as Class A and B(low risk) according to the classification rules for the GHTF guidance. Regulating condoms is justifiable since they are Class C devices for contraception or the prevention of sexually transmitted diseases (6). There is a need to adopt risk classification and conformity assessment proportional to the product’s risk class. This approach’s advantage is reduced workload and regulatory burden to the manufacturers. This, in turn, results in effective and efficient use of resources whilst making access to the products more quickly to the population that needs them the most (5).

On the other hand, the MLScCZ bases its registration only on a limited performance evaluation conducted at the NMRL. However, NMRL staff were not trained on the regulatory functions that require independent performance evaluation of IVDs to support market authorisation decisions. The NMRL did not have access to the Clinical and Laboratory Standards Institute (CLSI) that guides laboratorians in developing and implementing protocols for validating and verifying IVDs. NMRL did not have access to reference panels traceable to an International Standard. The reference panels are standardised specimens for calibration and harmonisation of assays that detect specific pathogens, for example, HIV antibodies or antigens and TB nucleic acids. Performance evaluation criteria were only based on diagnostic sensitivity and specificity characteristics. Other aspects include analytical sensitivity and specificity that challenge the product’s Limit of Detection (LoD), investigation of cross-reactivity, potentially interfering substances, precision and flex and robustness, and lot-to-lot variation of the product were not being assessed by the NMRL. The implications are that the MLCScCZ registers IVD medical devices and if not approved by any other stringent regulatory authorities, may not meet the safety and performance requirements to ensure the safety of the patients. The regulatory system must use reliance and recognition mechanisms to ensure that medical devices that may not have been assessed stringently by the regulatory authority in Zimbabwe but approved by other stringent regulatory authorities provide some assurance. However, the legislation in Zimbabwe does not allow reliance and recognition mechanisms. Reliance must operate within existing legal frameworks. This means that when an NRA relies on stringent assessments conducted by other jurisdictions, the relying NRA must still make its own decisions on approving or not approving the products. The relying authority remains independent, responsible and accountable regarding the decisions taken, even when it relies on the findings, assessments and information of others. The reliance mechanism is meant to reduce the barriers to access of IVDs due to unclear regulations and a lack of legal provisions to implement it. Regulatory reliance enables healthcare systems to;

- accelerate global access to safe and quality health technology,
- increase efficient use of resources and avoid duplication of efforts,
- reduce uncertainties for innovators and improve harmonisation in regulation,
- promote more consistent and robust responses to crises (15).

The WHO prequalification programme informs procurement of quality-assured IVDs in RLS. It uses a standardised procedure to assess if IVDs meet the specific safety, quality and performance requirements. The assessment includes a technical documentation review, manufacturing site inspection, performance evaluation and labelling review (16). It can also expedite the registration of IVDs in other countries through improved information sharing between WHO prequalification, NRAs and manufacturers through a reliance mechanism called the Collaborative Registration Procedure (CRP). Leveraging assessment and inspection outputs already produced by WHO prequalification, thereby eliminating duplicative regulatory work, speeds up in-country registration of quality-assured products.

Additionally, the CRP conducts assessments with RLS in mind. It contributes to their wider availability (17). This procedure can be one of the solutions for Zimbabwe to expedite IVD registration due to the limited capacity to regulate these products. Effective implementation of this mechanism may require the MASCA to be amended to include reliance mechanisms in the legislation.

Post-market surveillance (PMS) activities are not well documented. The MCAZ has not been doing much for condoms and gloves. "*Currently, we have not been doing it. We have not been doing much from our end. That has been our weakness. The only thing we have been doing is going around the country to check for unregistered medical devices. Our regulations are not so explicit on post-market surveillance, but that has been addressed in the draft regulations. Our strategy is that the pharmacovigilance team may handle it (MCAZ KII1)."* This contradicts Hubner et al.’s finding that the MCAZ had PMS in place (9). The MLCSCcZ acknowledged a process for pre-distribution lot-to-lot testing conducted at NMRL and a mechanism to report and investigate user feedback if potential failures of IVDs are suspected though not well documented. One respondent commented, "*So whenever they’re those adverse reports, the local distributor would immediately be informed and engaged with respect by the ministry of health through the lab directorate at that time although the things were not done correctly. But if the book followed them, those reports they have come out, and the technical advisory committee would have advised the lab director. This is why the council was involved; professionals regulated by the council are running these tests (MLCScCZ KII4."* Some contributing factors to such weaknesses could be a lack of capacity to establish, implement and maintain mechanisms for a robust PMS and a lack of effort in strengthening the regulatory system of medical devices, including IVDs. Dube-Mwedzi et al. also found these when they conducted a rapid assessment of national regulatory systems in SADC. This is consistent with the finding in the study conducted by Stanislav et al. that most of the countries in the SADC region did not have a policy framework to underpin the prevention, detection and response to incidences of substandard, falsified medical products, handling of market complaints, control of promotion of medical devices, detection of and action against substandard and falsified medical devices, removal and disposal of defective medical devices (18).

The interviewed participants recognised the need for collaboration and improving medical device regulation. “*Well, the need for regulation must be balanced, but we cannot do all the regulatory functions. There is a need to have the training and technical support, including infrastructure and equipment to conduct the regulatory functions, human resources that are trained and competent to conduct the work, and mechanisms to ensure transparency and accountability.”* (NMRL KII8). Another participant said, “*I think lack of consensus resulted in working in silos and on professional differences. Laboratory scientists look at IVDs as tools of their trade that must be regulated by scientists. This made it challenging for MCAZ and the MLCScCZ to work together, leading to a deadlock. Both institutions are on talking terms now, and we hope to see progress”* (MLCScCZ KII12). Therefore, when coming up with the road map, it is essential for the regulatory authority to consider different and applicable professionals to ensure that staff with the right qualifications, competencies and skills conduct the regulatory work. This requires a multi-sectoral approach using system thinking. The approach needs to consider the following key stakeholders;

1. Practitioners and researchers
2. Advocates
3. Industry representatives
4. Leaders.

Effective medical device regulation requires system organising, systems dynamics, systems networks and systems knowledge. The following are the definitions of the terms listed above;

- *“Systems organise to understand and foster the development of participatory, complex, and adaptive collaborative systems regulating medical devices; ensure effective facilitation and management; and encourage productive system action and learning*.
- *System dynamics to understand and model the complex dynamic interactions involved in the regulatory system and among the factors influencing medical device use, including political actions such as legislation, research advances, medical device control activities, industry forces, and social and cultural factors*.
- *System networks to understand and analyse effective collaborative relationships among stakeholders, improve collaboration strategies, and help reduce duplication of effort*.
- *Systems knowledge to develop and manage the knowledge infrastructure required for effective dissemination and evolution of scientifically credible, evidence-based practices, together with an effective strategy to package, deliver, and maintain this knowledge”* (figure 1) (19).

**Figure 1:**
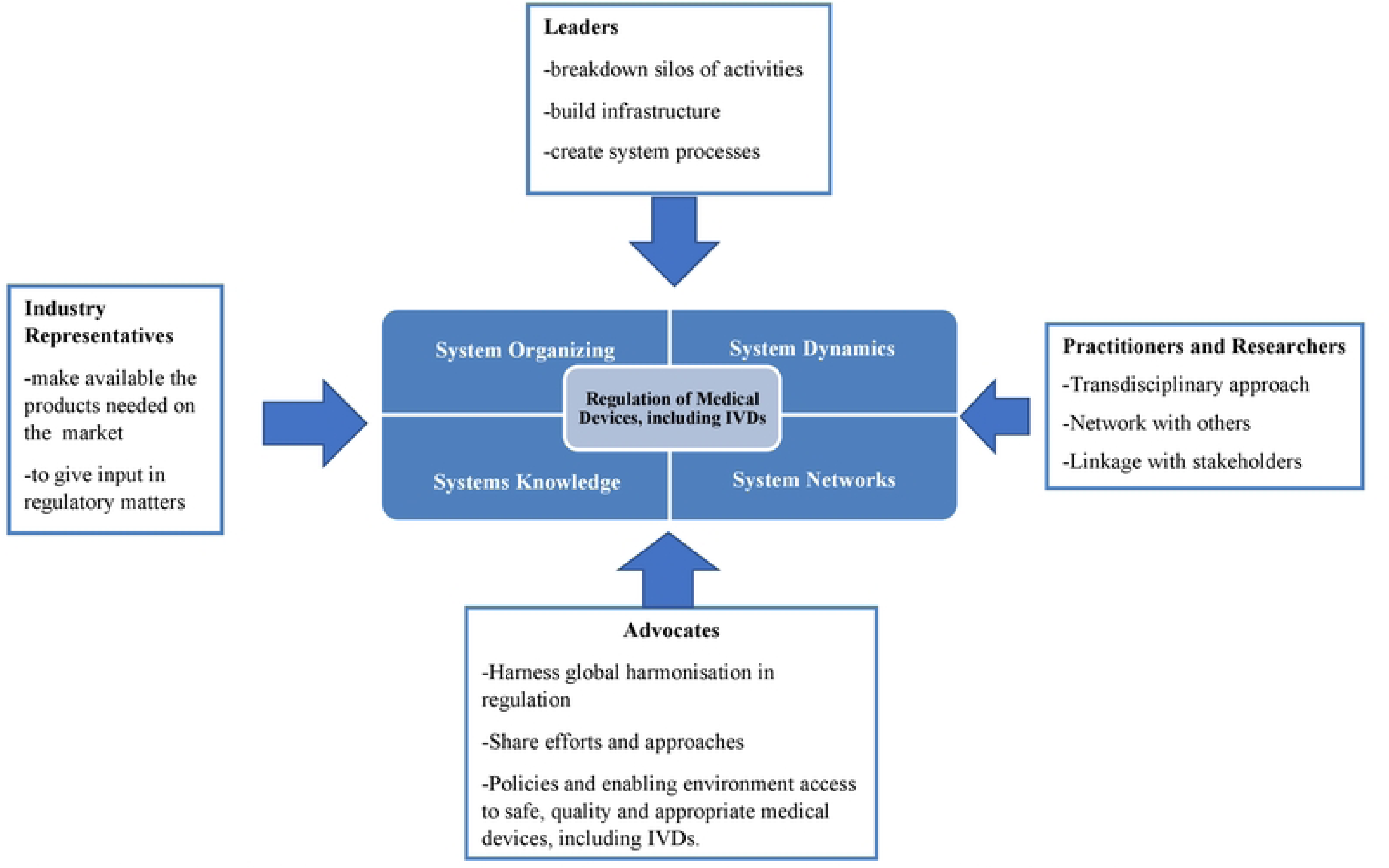
Proposed stakeholder engagement and systems thinking approach for medical devices regulation in Zimbabwe.

Concluding the findings from interviewing participants from Zimbabwe, it appears that Zimbabwe is on the pathway to strengthening medical device regulations. This requires collaboration and systems thinking to ensure the medical devices regulatory system is effective.

The GMRFMD is essential to guide NRAs to establish and strengthen the medical devices regulatory system. The findings have shown that the framework is popular among regulatory authorities in developing countries. Revision of the GMRFMD to include GBT + medical devices indicators into the GMRFMD. This revision intends to consider factors for advanced regulation of medical devices: "Understanding that this model was developed, uh, in 2016, the drafting was in 2016, publishing was in 2017, and years have passed since *this document was drafted and published. And so the discussion in the GBT, uh, indicators triggered revision.”* (WHO KII13). South Africa modelled its medical devices regulatory systems from the GMRFMD and the WHO Global Benchmarking Tool. "*So the roadmap indicated that they’re following the WHO Global Benchmarking (GBT) process for implementing the medical device regulations in the country (SAHPRA KII14)".* SAHPRA and TMDA had to develop an action plan to improve the regulatory system of medical devices. In the case of South Africa, it had to benchmark the medical devices regulatory system using the GBT to determine the baseline status.

Additionally, the regulator compared its regulatory approaches with other agencies, namely Australia’s Therapeutic Goods Administration (TGA), Canada’s Health Canada, Singapore’s Health Science Authority (HSA) and Switzerland’s Swissmedic. The purpose of the comparison was to identify areas of improvement and harmonise the regulatory system. The study also informed the re-engineering of the NRA, resulting in its name change from the Medicines Control Council (MCC) to the South African Health Products Regulatory Authority (SAHPRA). It was concluded that MCC used similar Good Regulatory Review Practices (GRevP) compared to Australian, Canadian, Singapore, and Swiss NRAs. However, review timelines to grant product market authorisation were more extended than other NRAs (20,21). Similarly, Tanzanian NRA built its case by comparing the then position and best practices from USFDA, TGA and PMDA: *“So we made sure that we had a, a lot of references, we had a lot of information from other mature jurisdictions such as, such as the USFDA, TGA, Japan, all these other big, uh, national regulatory authorities to say what is happening in other countries that have made, uh, uh, tremendous, uh, progress in this, in this space for harmonisation purposes.”* (WHO KII15). Another study also included Health Canada to benchmark the medical devices regulatory authority (12). The advantage of this approach is that it also was to harmonise the regulations to implement reliance and recognition mechanisms. Stakeholder engagement and advocacy are essential to capture all the stakeholders’ views. Stakeholders must include policymakers, regulators, and economic operators to ensure acceptance of the regulations.

There is a need to promote convergence and harmonisation of medical device regulations. Convergence is a process *"whereby the regulatory requirements across countries or regions become more similar or "aligned" over time as a result of the gradual adoption of internationally recognised technical guidance documents, standards and scientific principles, common or similar practices and procedures, or adoption of regulatory mechanisms that might be specific to a local legal context but that align with shared principles to achieve a common public health goal." Harmonisation is a process by which technical guidelines are developed to be uniform across participating authorities.* (22). Learning from the South African and Tanzanian experiences, convergence, and harmonisation are vital in reducing the regulatory burden on manufacturers and regulators. Furthermore, in the study conducted by Keyter et al., it was recommended to implement facilitated regulatory pathways and apply a risk-based approach to the regulatory review process to conserve limited resources, avoid duplication of regulatory efforts and get the medical devices to the populations that need them the most (20). The experiences from South Africa and Tanzania are essential to Zimbabwe amending the MASCA to address the following;

- *"Explicitly state the institution (s) with the legal mandate to regulate medical devices with powers to enforce the regulations and market oversight*.
- *Define medical devices and the scope of medical devices regulated under this law, ensuring the regulatory framework is capable of adapting to new technologies and treatment modalities*.
- *Stakeholder responsibilities, powers to issue implementing regulations and to take action where the health of patients or users is compromised, and the responsibility for publishing guidance documents to aid understanding of legal requirements;*
- *provide the regulatory authority with administrative and enforcement discretion for reliance upon and recognition of the work or decisions of regulatory authorities in other jurisdictions;*
- *require that only safe medical devices that perform as the manufacturer describes in its labelling may be placed on the market;*
- *specify market entry conditions for medical devices;*
- *establish record-keeping, registration and reporting requirements for all parties within the scope of the law, including the regulatory authority;*
- *specify a transition period sufficient to allow parties affected by the law to comply with its requirements and ensure minimal disruption to the continuing supply of medical devices to health facilities and other users"* (13).

The challenges encountered in the transition of the regulations were the capacity and competencies to regulate medical devices, including IVDs. Most personnel within TMDA were pharmacists and were unfamiliar with frameworks for medical devices, including IVDs. There was the need for a complete mindset shift during the transition period: to switch from pharmaceuticals to medical devices and regulation. The interviewee commented, "*I say that, of course, you must learn. So when it comes to learning, you need people willing to learn and consider that. And of course, medical devices are diverse, so it depends on multi-disciplinary teams for the issues and how to accommodate the groups”* (TMDA KI15). In the context of SAHPRA, the interviewee said, “*The structure comprised chiefly of the pharmacists needed some changes to accommodate other professionals, such as medical engineers and scientists, to advice on regulatory affairs of medical devices* (SAHPRA KI14)." The TMDA now has a multi-disciplinary team of external experts associated with the design, development, manufacturing, evaluation and use of medical devices that was put in place to advise the TMDA with basic understanding and conformity assessment-related work, including all other regulatory functions of the NRA. The medical devices department has now grown to an entire directorate within the TMDA, with about 40 staff members employed by the TMDA. The other challenge the TMDA faced was the temptation to use the medicines regulatory framework on medical devices, including IVDs. It is appreciated that the medicines regulatory framework has matured over the years and is stable, but it does not apply to medical devices. The interviewee commented, *"For example, in the context of medicines, it is apparent that from production, products with a shelf life of 36 months should have at least 80% of the shelf life remaining. Medicines with a shelf life of 24 months should have at least 60% remaining by the time they arrive at the port of entry (TMDA KII15)."* This approach does not apply to reagents and controls that fall under IVDs with a shorter shelf life of fewer than six weeks; by the time they arrive at the port of entry, they might be left with two weeks. The interviewee commented, *"So in terms of when you start regulations, it’s imperative to disengage regulations from those medical devices, from the ones for medicine, as early as possible, to make people open-minded, and to treat medical devices the way they are supposed to be treated following their life cycle* (TMDA KII15)." Therefore, it is recommended that the NRA develop a framework specific for medical devices following medical devices’ lifecycle depending on the nature of design and intended use of the medical device.

The main output of this study is a roadmap for Zimbabwe to establish, implement and continuously improve a responsive, predictable, standardised and transparent regulatory system oriented to the outcome. The proposed roadmap uses a tired approach. The three tiers are legislation, regulations and guidance. Legislation subjected to public scrutiny and does not contradict the Constitution is the mainstay of regulations. Regulations interpret the law and make the implementation of the law feasible. The guidance provides manufacturers with instructions to meet the requirements of the regulations (2).

Furthermore, a competence framework is proposed because human resources are important for an effective medical regulatory system. The competence framework is adopted from the Regulatory Affairs Professionals Society competence framework and is cross-cutting on legislation, regulations and guidance (23). The proposed road map is shown in Figure 2 below.

**Figure 2:**
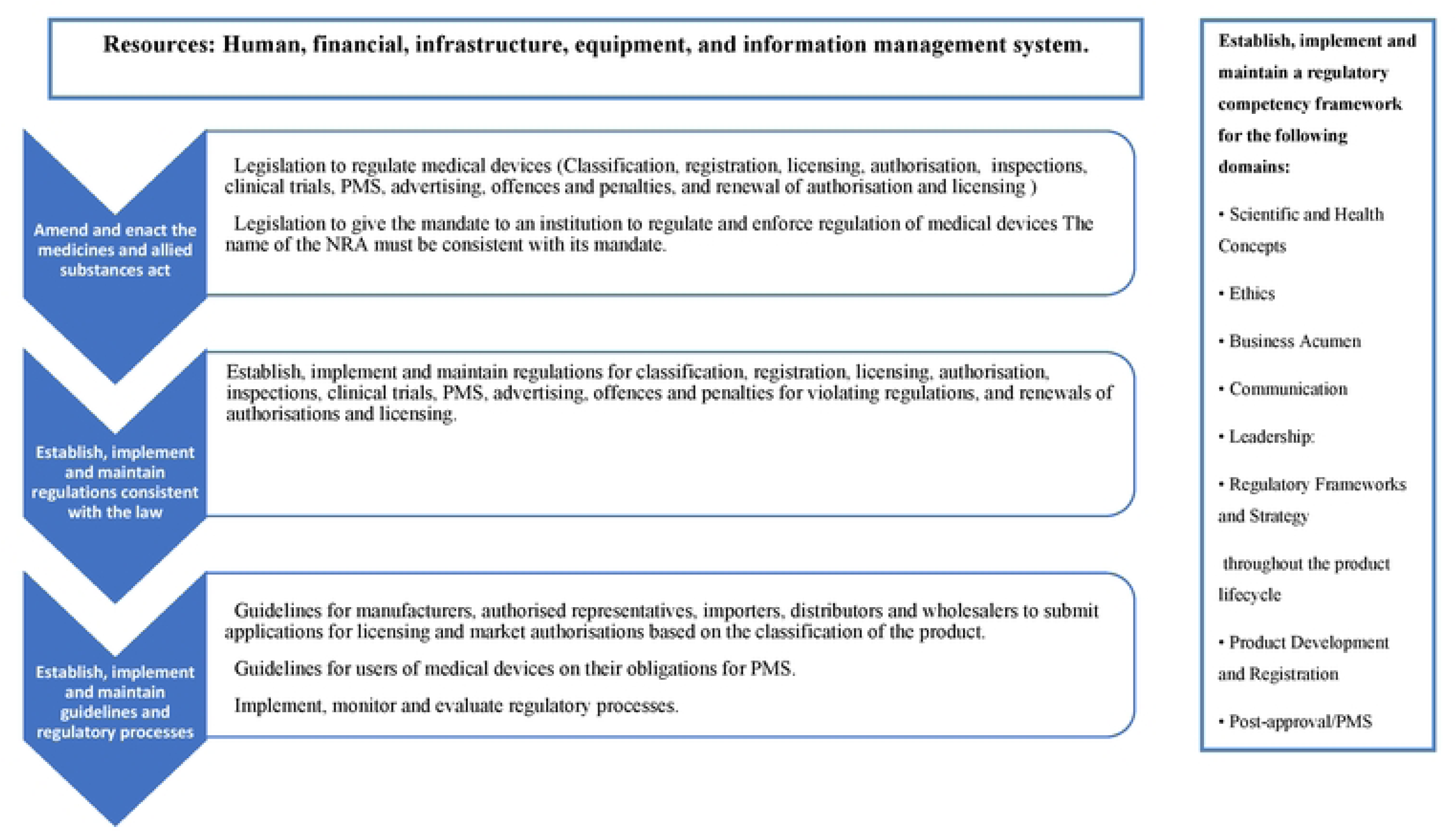
A road map to address deficiencies in medical devices regulation in Zimbabwe.

### Study Limitations

There are limitations to this research, as well as issues related to implementing a case study. One of the limitations is that the study did not include the Ministry of Health officials, users of the medical devices, the donor community, procurers, manufacturers, importers, authorised representatives and distributors of medical devices to get their perception of the current state of medical devices regulation in Zimbabwe. The results of this study cannot be generalised to the broader research community due to the nature of the research strategy. The researcher uses a tried and tested research strategy, appealing to relatability rather than generalizability. However, it was also argued that generalisation, although not immediate, can take place over some time – incremental generalizability – as more empirical research case studies are implemented. Data saturation was not achieved regarding sample size as the number of interviewees from Zimbabwe did not reach 15.

## Conclusions

Medical devices in Zimbabwe are not adequately regulated. The role of effective medical device regulation is recognised to ensure that safe medical devices of acceptable quality and performance are made accessible to the populations that need them the most without delay due to unclear regulatory processes. The legal framework needs to be amended to ensure that medical device regulation is founded on a sound basis in law. It clearly states the institution(s) with the legal mandate to regulate and enforce the regulations, define the products within its scope, and identify the entities subject to regulation. The formulation, implementation and enforcement of medical device regulations require systems thinking and multi-sectoral collaboration. Harmonisation and reliance practices will make the regulatory system predictable and proportional to the risk classification of medical devices and reduce the regulatory burden on economic operators and the workload on the regulatory authority. Hence, it will make medical devices expeditiously accessible.

## Data Availability

All relevant data are within the manuscript and its Supporting Information files.

## Acknowledgements

The authors would like to acknowledge the cooperation of the agencies and personnel interviewed in Zimbabwe, South Africa, Tanzania, and the WHO and for their assistance with accessing documentary evidence and reports.

